# Trends in inequalities in avoidable hospitalisations across the COVID-19 pandemic: A cohort study of 23.5 million people in England

**DOI:** 10.1101/2022.12.14.22283458

**Authors:** Mark A. Green, Martin McKee, Jon Massey, Brian MacKenna, Amir Mehrkar, Sebastian Bacon, John Macleod, Aziz Sheikh, Syed Ahmar Shah, The OpenSAFELY Consortium, The LH&W NCS Collaborative, Srinivasa Vittal Katikireddi

## Abstract

**Background:** The COVID-19 pandemic and associated national lockdowns created unprecedented disruption to healthcare, with reduced access to services and planned clinical encounters postponed or cancelled. It was widely anticipated that failure to obtain timely treatment would cause progression of illness and increased hospital admissions. Additional concerns were that social and spatial inequalities would widen given the disproportionate impacts of COVID-19 directly. The aim of our study is to determine whether this was observable in England.

**Methods:** With the approval of NHS England we utilised individual-level electronic health records from OpenSAFELY, which covered ∼40% of general practices in England (mean monthly population size 23.5 million people). We estimated crude and directly age-standardised rates for potentially preventable unplanned hospital admissions: ambulatory care sensitive conditions and urgent emergency sensitive conditions. We considered how trends in these outcomes varied by three measures of social and spatial inequality: neighbourhood socioeconomic deprivation, ethnicity, and geographical region.

**Findings:** There were large declines in avoidable hospitalisations during the first national lockdown, which then reversed post-lockdown albeit never reaching pre-pandemic levels. While trends were consistent by each measure of inequality, absolute levels of inequalities narrowed throughout 2020 (especially during the first national lockdown) and remained lower than pre-pandemic trends. While the scale of inequalities remained similar into 2021 for deprivation and ethnicity, we found evidence of widening absolute and relative inequalities by geographic region in 2021 and 2022.

**Interpretation:** The anticipation that healthcare disruption from the COVID-19 pandemic and lockdowns would result in more (avoidable) hospitalisations and widening social inequalities was wrong. However, the recent growing gap between geographic regions suggests that the effects of the pandemic has reinforced spatial inequalities.

## Introduction

There has been widespread public, political, and media concern that the unprecedented disruption in access to care associated with the pandemic would have severe consequences for population health beyond the immediate effects of COVID-19. During the first UK national lockdown, much NHS activity was postponed or cancelled. There was some catching up in the summer of 2020 as capacity allowed, but subsequent waves of infection accompanied by lockdowns have had similar impacts. While the NHS has adapted, with new ways of working and physical reconfiguration within hospitals, the need for enhanced infection control measures coupled with high levels of staff illness from both acute COVID-19 and, increasingly, Long COVID, have caused significant disruption that continues to affect the level of care delivered.^1^ This has impacted trends in health system performance. Visits to general practice fell markedly,^2^ although this was later compensated for by a rise in online consultations.^3^ There were three million fewer elective treatment pathways in England in 2020 than compared to 2019.^4^ Cancer screening programmes, non-essential surgeries and diagnostic procedures were postponed or cancelled.^2,5^ Waiting lists have continued to get longer, resulting in delayed access to care for new treatments.^6,7^ The comprehensive disruption to multiple pathways across the health system, encompassing prevention, treatment and management was therefore predicted by many to lead to an increase in hospital admissions as a result of people not receiving the care they needed.

There was a particular concern that this disruption would widen existing inequalities. The COVID-19 pandemic shone a light on the fractures within English society,^8^ with the poor, ethnic minorities, and those living in parts of northern England disproportionally affected.^9–11^ Early evidence suggests that disruption of elective treatment pathways has been greater in deprived areas.^4^ Understanding whether differential experiences of healthcare disruption resulted in greater hospital admissions will be essential if we are serious about reversing the inequalities that have been exacerbated by the pandemic. Narrowing health inequalities, especially in hospital admissions, is a government priority.^12,13^

Identifying the impacts of healthcare disruption is difficult. The experience of those experiencing illness can vary considerably. For example, it is widely accepted that, despite concerted efforts over many decades, the inverse care law persists.^14^ The journey from onset of illness is complex and obstacles can arise at many points.^15^ To simplify this issue, we borrow a key concept from the health systems literature, ‘avoidable hospital admissions’, considered a proxy measure for health system performance.^16^ Avoidable hospitalisations are emergency (unplanned) hospital admissions that could have potentially been prevented if individuals had received timely care within the community (and may be susceptible to disruptions to care).^16,17^ Reducing avoidable hospital admissions is a priority for the NHS because they are often costly and disrupt elective care.^12^ We hypothesised that the significant disruption to healthcare access in the community associated with uncontrolled infection and non-pharmaceutical interventions implemented in response (e.g., people unable to see their GP or postponement to treatments) may have resulted in more avoidable hospitalisations since individuals were not able to receive the care they needed.

Our aim is to describe trends in avoidable hospitalisations during different periods of the COVID-19 pandemic and understand how these were affected by measures of social and spatial inequality.

## Methods

### Data

The primary data source for the analysis was OpenSAFELY-TPP. OpenSAFELY is an open-source secure health data analysis platform providing access to primary care records, linked to secondary care and mortality records, held by the two largest electronic health record providers for NHS England – Egton Medical Information Systems (EMIS) and The Phoenix Partnership (TPP) which cover ∼58 million registered patients. In this study, we had access to data from TPP (The Phoenix Partnership) which covers ∼40% of general practices in England. The data are broadly representative of England by age, sex, ethnicity, small area socioeconomic deprivation and cause of death.^18^

Data were accessed on 20^th^ May 2022. The study period for our analysis was 1^st^ January 2019 to 31^st^ March 2022. While 2019 only provides one year’s worth of pre-pandemic data, trends prior to 2019 were relatively flat.^19^ We discounted all data in April and May 2022 to minimise under-counting of events due to possible delays in reporting of clinical information relating to admissions. Total counts per month were rounded to their nearest 5 to minimise patient disclosure risks where low counts were evident. All analytical scripts, processed data and outputs are openly available at https://github.com/opensafely/avoidable_hospitalisations_trends.

### Outcomes

Avoidable hospitalisations were defined using five measures commonly used by NHS England.^16^ Each were defined using ICD-10 codes (codelists openly available^20^), primary diagnosis (diagnostic position 1) and for emergency (unplanned) admissions only. All ambulatory care sensitive conditions were selected as an overall proxy for avoidable hospitalisations. Ambulatory care sensitive conditions were defined as conditions that can be treated effectively in the community and should not therefore require hospital admission.^16,17,19^ We further disaggregated all ambulatory care sensitive conditions into three measures representing the type of condition: (i) acute conditions (e.g., cellulitis, dental caries, rickets, gastric ulcer), (ii) chronic conditions (e.g., hypertension, angina, asthma), (iii) vaccine-preventable conditions (e.g., mumps, measles, influenza). We did not include COVID-19 admissions as avoidable or vaccine-preventable conditions as we wanted to examine the indirect impacts of the pandemic (as well as ensuring a consistent set of conditions pre-pandemic). We also included emergency urgent care sensitive conditions as an alternative measure of avoidable hospitalisations. These are acute exacerbations of urgent conditions that will result in hospital admission but that the NHS should be able to treat within the community to minimise the need for hospital care.^16,17,19^ Finally, we also include a measure of all emergency hospital admissions to provide context for our measures.

### Exposures – measures of inequality

We considered how trends in our outcome variables varied by three measures of social and spatial inequality that have been widely reported to have been associated with unequal COVID-19 outcomes.^8,13^ Neighbourhood socioeconomic deprivation was measured using the Index of Multiple Deprivation 2019 (IMD).^21^ Individuals were matched to Lower Super Output Area by their home residence in each month and we calculated the quintile of deprivation rank. Ethnicity was recorded in the electronic health record and we consider four groups (White or White British, Black or Black British, Asian or Asian British, and Mixed ethnicity). We did not undertake further disaggregation because of issues with small numbers. We excluded individuals with ‘Other’ ethnicity since the group is extremely heterogeneous, limiting our scope to draw conclusions (they represented 2.2% of the entire sample). Patients were allocated to seven Government Office Regions, based on home residence, to capture regional differences. Missing data for covariates are presented in Appendix Table A.

### Statistical analyses

We estimated summary statistics for each outcome measure using aggregated counts from individual records. Where aggregated statistics were calculated, we used crude rates to measure monthly hospital admission rates (population was defined as total number of people alive on the 1^st^ of each month within the OpenSAFELY-TPP data). Where we stratified measures by socio-demographic indicators, we estimated directly age-standardised rates to adjust for the different age structures of each group that may confound trends. All measures were stratified by sex (female or male). We visualised rates and presented descriptive statistics to investigate trends. To aid the interpretation of trends, we also calculated indicators to summarise the extent of disparities for each month. For deprivation, we estimated the slope index of inequality (SII) which provides an estimate of the absolute level of inequality by neighbourhood deprivation, incorporating information across all categories.^22^ We also estimate the relative index of inequality (RII) to describe the relative differences across categories. We were unable to calculates SII and RII for ethnicity and region as there is no ordinal structure to these variables. Here we estimated the range of values to describe the absolute level of inequality and the ratio of the maximum and minimum values to describe the relative level of inequality. We excluded all missing data from analyses.

## Results

### Overall population-level trends

Table 1 presents key summary statistics for our data. During the study period, there were 6 645 550 emergency hospital admissions, with a monthly average of 170 399. There were 1 129 770 ambulatory care sensitive hospital admissions (17% of all emergency admissions), chronic ambulatory care hospital admissions being most common. There 1 031 205 emergency urgent care hospital admissions (16% of all emergency admissions).

**Table 1:** Frequency counts for outcome measures.

| Measure | Total over study period | Mean per month |
| --- | --- | --- |
| Emergency admissions | 6,645,550 | 170,399 |
| All ambulatory care admissions | 1,129,770 | 28,968 |
| Acute ambulatory care admissions | 292,630 | 7,503 |
| Chronic ambulatory care admissions | 563,740 | 14,455 |
| Vaccine-preventable ambulatory care admissions | 282,630 | 7,247 |
| Emergency urgent care sensitive | 1,031,205 | 26,441 |
| Population | 918,302,035 | 23,546,206 |

Figure 1 presents crude admission rates by sex over the study period for each of our outcome measures. Trends in emergency hospital admissions were stable and consistent throughout 2019 (Figure 1A). In 2020, there were sudden large falls in emergency admissions coinciding with the first national lockdown in England (e.g., in April 2020 rates were 44% lower (females) and 39% lower (males) than compared to rates in January 2020). Emergency admission rates then increased quickly in the period following the national lockdown, although increases did not reach the same levels as in 2019. Rates fell once more at the end of 2020 (again coinciding with national lockdowns), subsequently recovering quickly and remaining at a level higher in 2021 than in 2020, but still less than in 2019.

**Figure 1:**
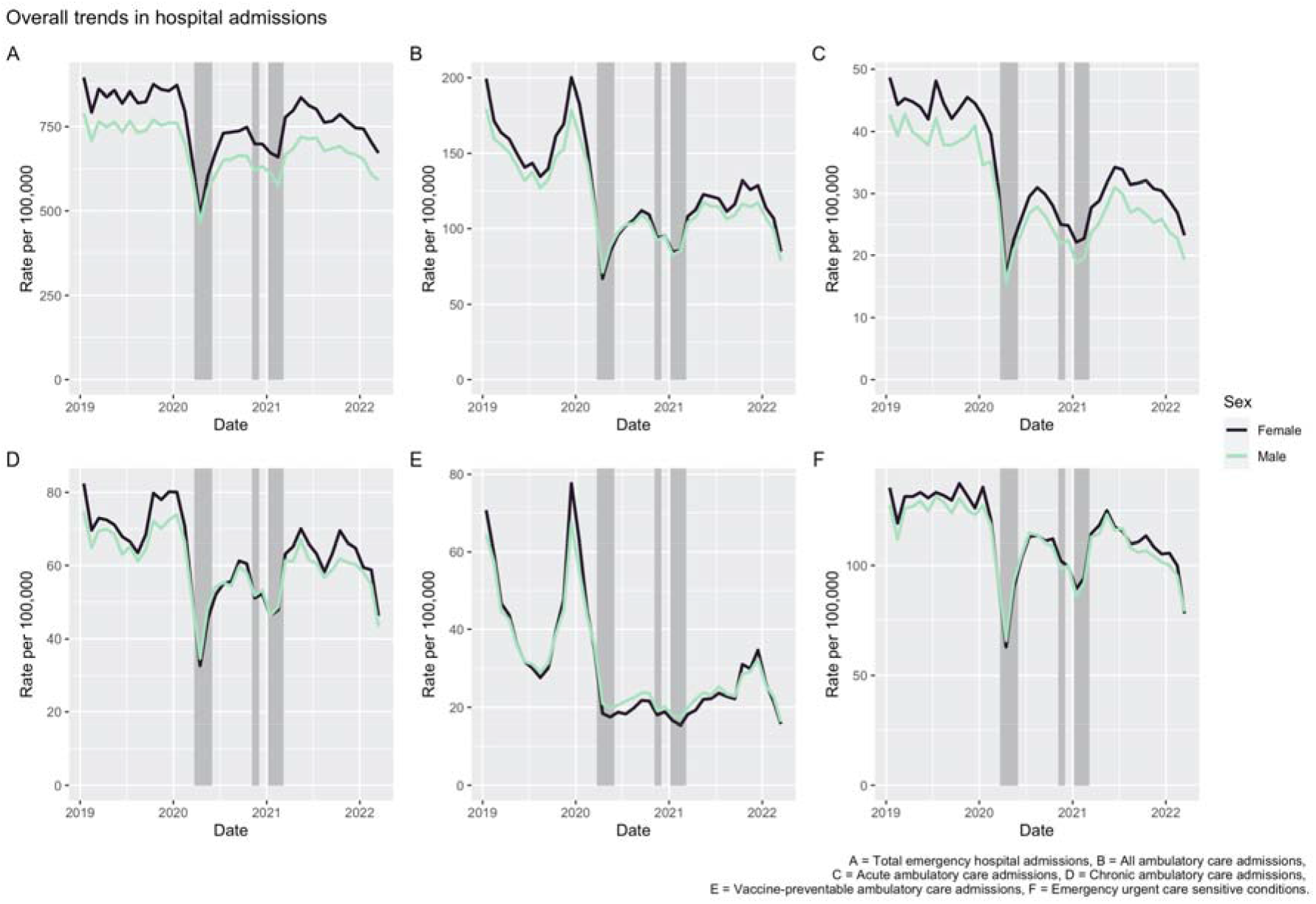
Crude hospital admission rates (per 100,000 population) by sex for measures of avoidable hospitalisations. Note: shaded periods represent national lockdowns.

Patterns of avoidable hospitalisations largely follow similar trends to emergency hospital admissions. There were large falls in all, acute, and chronic ambulatory sensitive hospital admissions and emergency urgent sensitive admissions between March and May 2020. These falls were then followed by large rises, albeit not to 2019 levels, before subsequent falls during the second and third national lockdowns (with large rises following). Vaccine-preventable ambulatory hospital admissions were the only measure that did not follow this trend. There were distinct peaks in winter 2019 but, after December, rates fell sharply and then continued at a low level throughout all of 2020. Rates then began to rise throughout 2021 (although only to the lowest points of 2019), peaking in December 2021, before falling thereafter.

### Trends by deprivation

We next examined whether trends in our measures varied by deprivation. Appendix Figures A and B present trends for females and males respectively, although there were minimal differences by sex so we describe the findings together. A distinct social gradient was evident for all measures, with the highest rates in the most deprived quintiles. Trends largely follow those reported at the population level and appear consistent across deprivation quintiles.

To aid the interpretation of these trends, we calculated the SII to provide an estimate of the absolute level of inequality (Figure 2). During the periods when rates fell sharply overall, SII values also did, indicating that absolute inequalities narrowed during these periods. While inequalities increased again when admission rates increased overall, the SII values did not increase to the same value as in 2019. The SII value appears to have continued to improve (i.e., narrowing inequalities) since late 2021 as well. Trends in relative inequalities (RII) were more erratic and displayed no discernible trends (Appendix Figure C).

**Figure 2:**
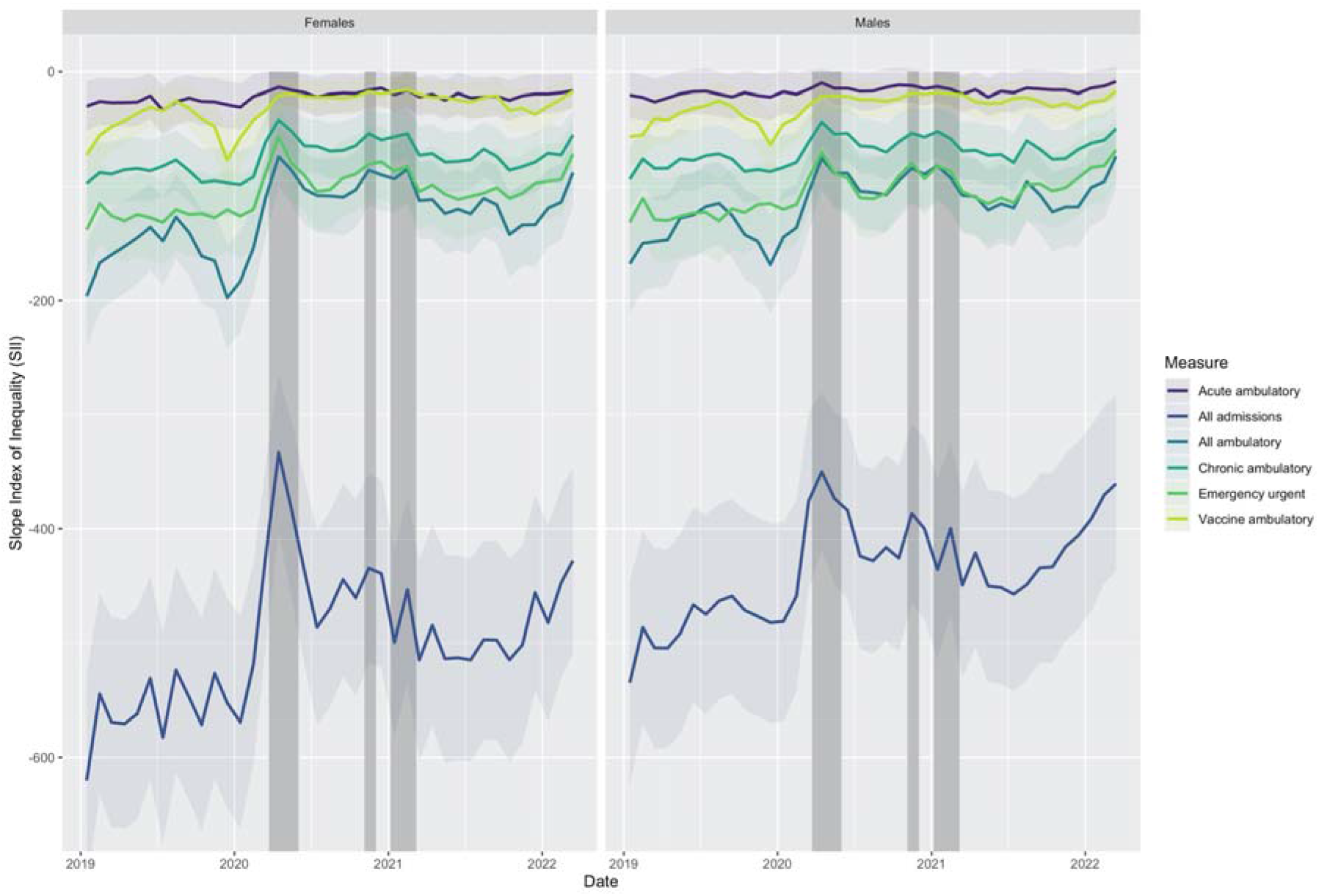
Estimated Slope Index of Inequality (SII) for socioeconomic deprivation by sex.

### Trends by ethnicity

Appendix Figures D and E present trends by ethnic group and sex. The general trends for each outcome match those described in earlier sections, with the falls in hospital admissions during key COVID-19 waves consistent across all ethnic groups. While there is some ordering of rates by ethnic group, the ordering itself is not consistent and confidence intervals often overlap, suggesting that any ordering is not meaningful.

We estimate the range (maximum value minus minimum value) for rates as an estimate of the extent of absolute inequalities (Figure 3). For all outcomes other than acute ambulatory admissions in females, absolute levels of ethnic inequalities have narrowed over time and especially post-pandemic. There was no clear relationship with the main waves/lockdowns for these measures. When considering relative inequalities, trends were erratic and inconsistent across outcomes (Appendix Figure F). Relative inequalities have broadly remained similar with no clear trend for chronic ambulatory and emergency urgent conditions (albeit with narrowing of inequalities during 2020 in females). Relative inequalities for acute ambulatory conditions have increased over the study period. Trends in all emergency admissions and all ambulatory admissions have narrowed over the period.

**Figure 3:**
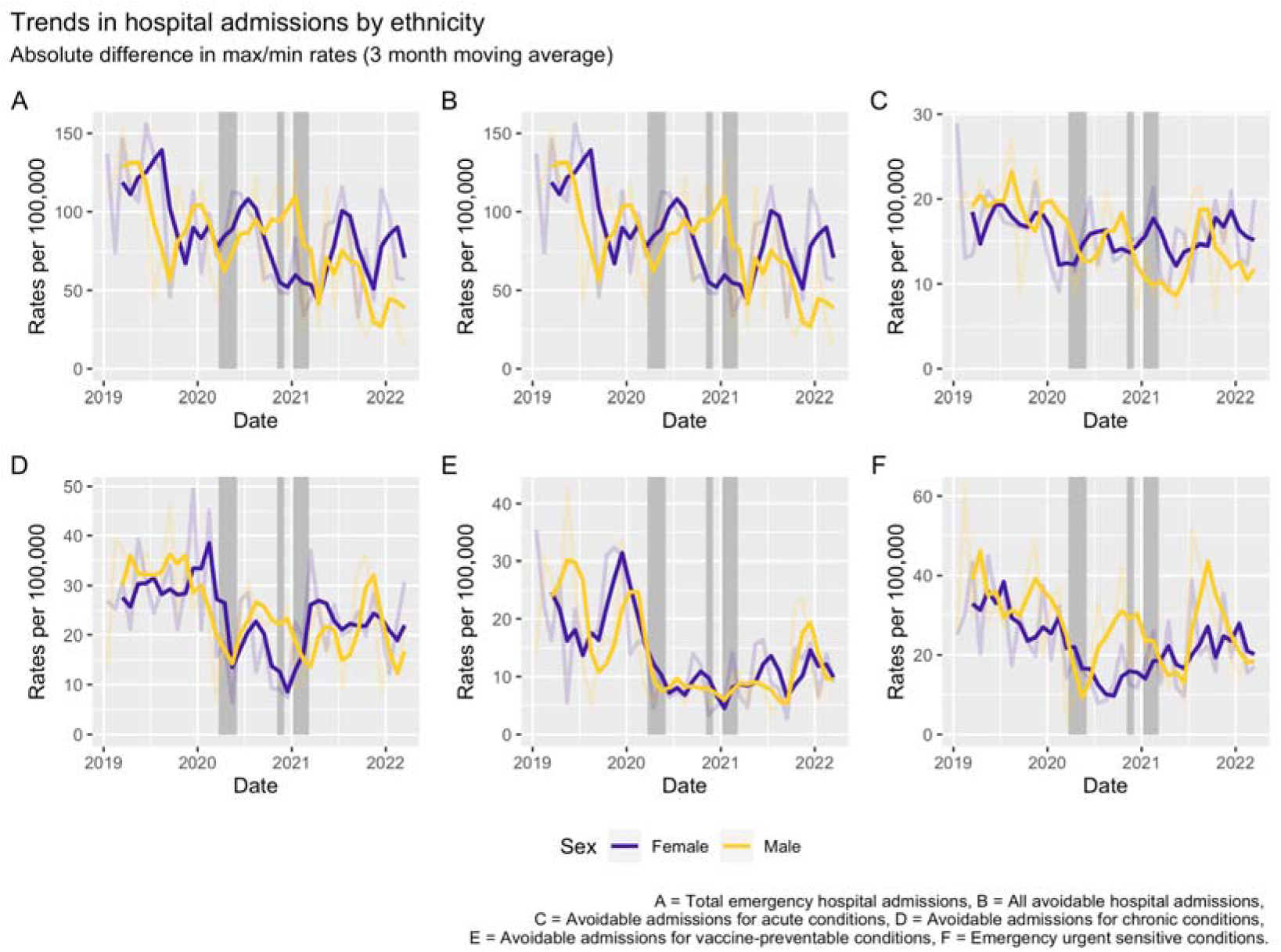
Absolute differences in directly age-standardised rates for admission (per 100,000) by cause between the maximum and minimum values across ethnic groups.

### Trends by region of England

Appendix Figures G and H present trends by geographical region for England. Trends largely follow those described previously, with falls in admissions across all outcomes during national lockdowns consistent across all regions. There are noticeable inequalities between regions. Rates in the West Midlands were consistently highest for most outcomes (other than vaccine-preventable admissions), followed by North West and North East. Admission rates in the South East and South West were consistently lowest across all periods and for all outcomes. London is the exception, lying in the middle for most outcomes. However, during the first national lockdown (and to a lesser extent at the end of 2020), falling rates take London to the lowest values.

We observe declining levels of absolute inequalities among regions (Figure 4) between 2019 and 2021. The falls are, mostly, greatest during national lockdowns, the only exception being vaccine-preventable ambulatory conditions, where the gap widened considerably during winters. From 2021 onwards, we observed widening inequalities across all of our outcomes. These trends are largely consistent by sex.

**Figure 4:**
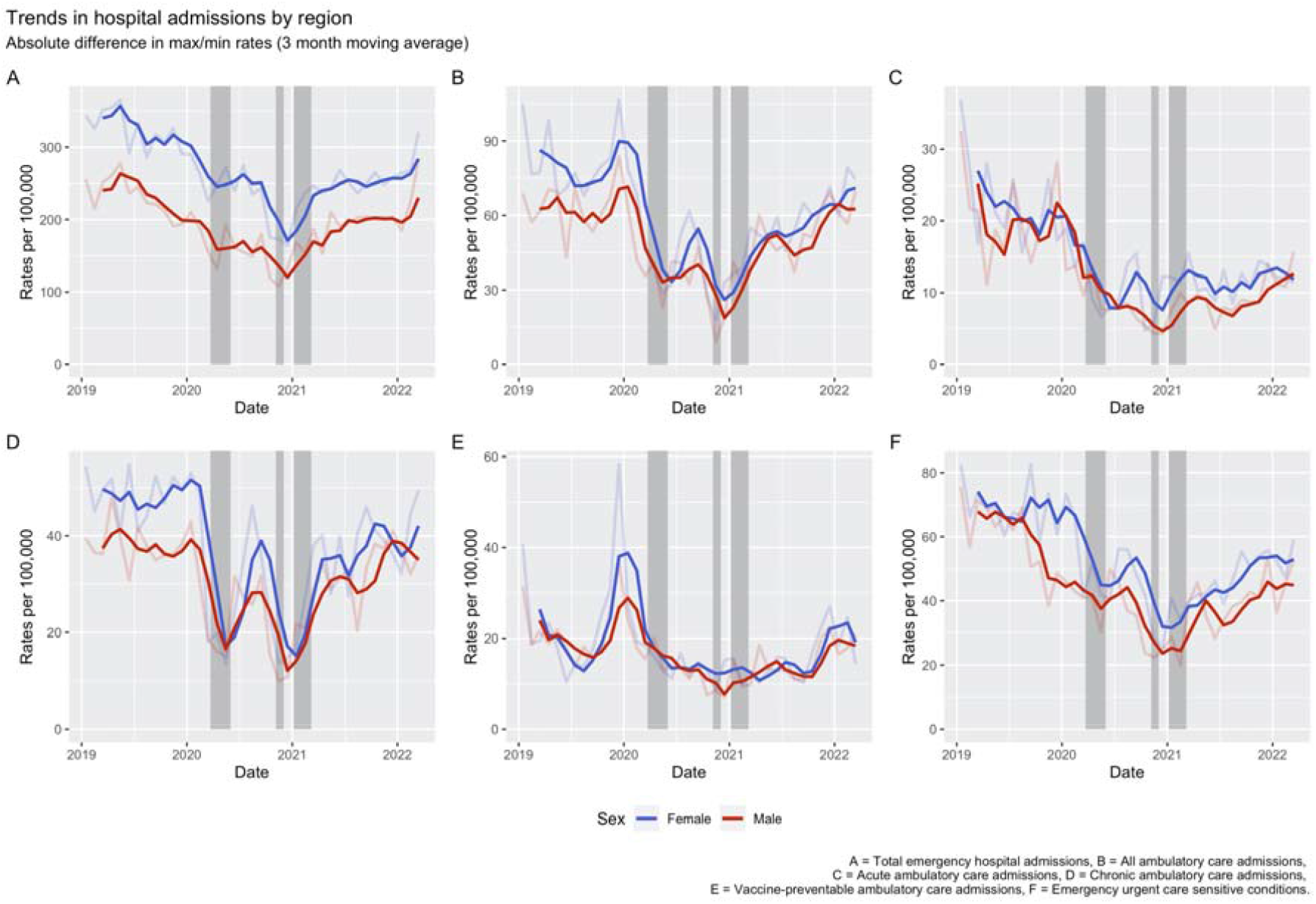
Absolute differences in directly age-standardised rates for admission (per 100,000) by cause between the maximum and minimum values across regions.

Turning to relative inequalities (Figure 5), in 2019 trends in our avoidable hospitalisation measures were flat. For all emergency admissions, we observe a decline in relative inequalities suggesting that overall gains in narrowing of inequalities in emergency admissions were not translated into narrowing avoidable hospitalisations. During the first COVID-19 wave (and national lockdown), we see large increases in relative inequalities across regions for all our measures other than chronic ambulatory admissions. Relative inequalities then decline subsequently through 2020. By the second and third lockdowns, trends for relative inequalities by regions were inconsistent across the measures. Relative inequalities widened for acute and vaccine-preventable ambulatory conditions, and emergency urgent care sensitive conditions, but narrowed for all emergency admissions, chronic and all ambulatory admissions. For all outcomes, we observe increasing relative inequalities for regions during later 2021 and into 2022.

**Figure 5:**
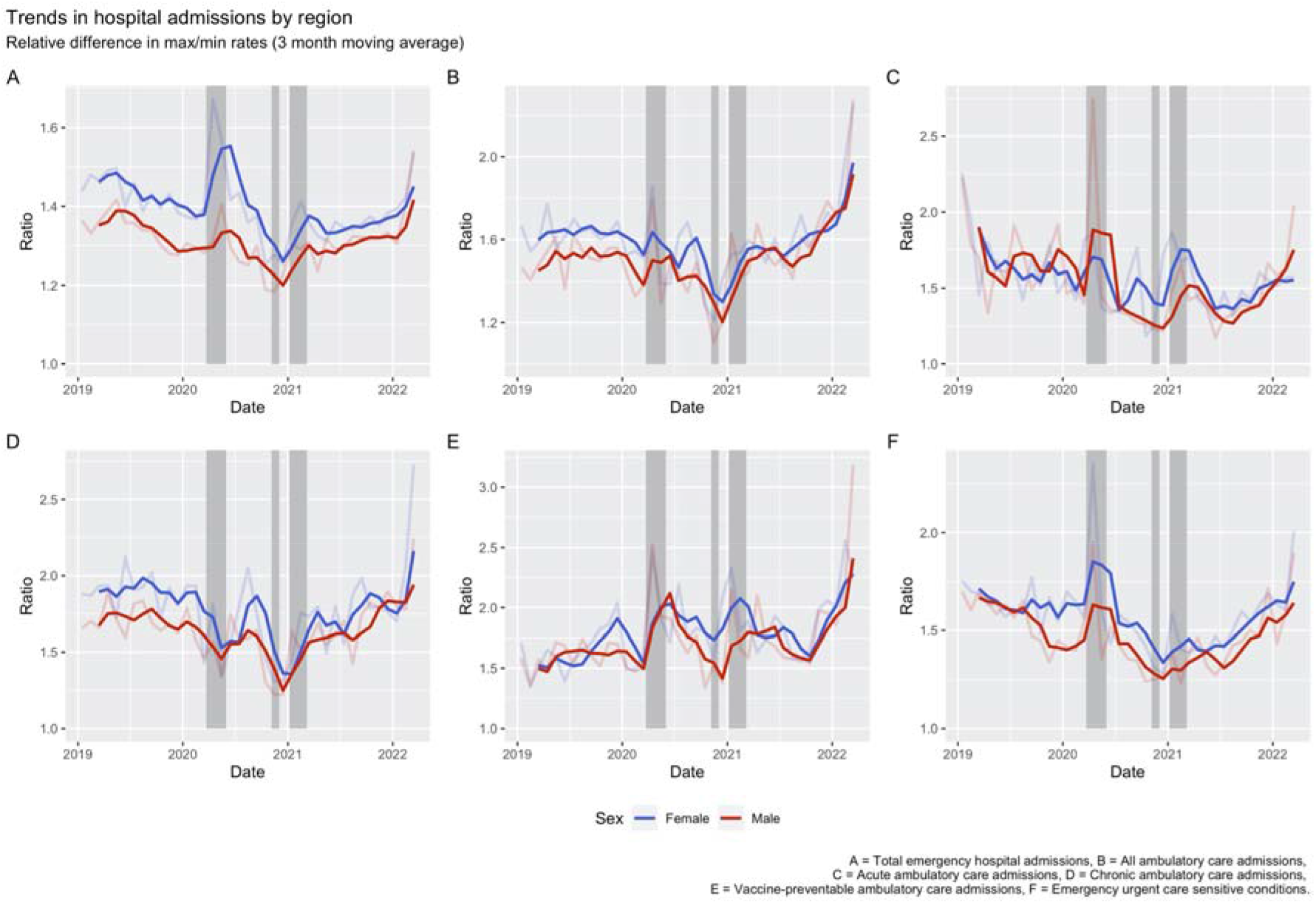
The ratio of directly age-standardised rates for admission (per 100,000) between the maximum and minimum values across regions and by cause.

## Discussion

### Key results

Many commentators anticipated that we would see increased hospitalisations as a result of people being disrupted in accessing the care they needed. Our study suggests that such fears were unfounded. All measures of avoidable hospitalisations we analysed declined during the first national lockdown. While rates largely rebounded subsequently (other than in periods of national lockdowns), avoidable hospitalisations have remained lower than 2019 levels. Trends in avoidable hospitalisations also fell in early 2022. We find evidence of narrowing absolute levels of inequalities by deprivation, ethnicity, and geographical region. The largest falls in inequalities occurred during the first national lockdown and have remained lower thereafter. During later parts of 2021 and into 2022, we find evidence of widening absolute and relative inequalities for geographical region only. It suggests that the recovery from the pandemic has reinforced spatial, more than social, inequalities.

### Interpretation

Our study is one of the first and largest to investigate how avoidable hospitalisations changed before and during the COVID-19 pandemic. While we originally hypothesised that we might expect higher avoidable hospital admissions post pandemic and during national lockdowns, our results suggest the opposite. Thus, we find large falls in avoidable hospitalisations during periods of national lockdowns. During periods when society opened up, rates increased but never to 2019 levels suggesting a gradual recovery in population-level trends in avoidable hospitalisations. Our results follow observations for other metrics including falls in non-COVID-19 mortality.^23,24^ Put simply, what everyone anticipated would happen did not and we need to know why this was.

While one interpretation might therefore be that the impacts of healthcare disruption were minimal, we should not forget that healthcare disruption is part of the explanation for the trends we observe. During the first national lockdown, the NHS cancelled or postponed or many patients planning to be admitted to free capacity for patients with COVID-19.^4,5,7^ Individuals may also have avoided going to hospital if at all possible for fear of being exposed to SARS-CoV-2.^25^ Such disruption would explain why we saw the largest decreases in rates during the first lockdown; the NHS was trying to avoid admitting people to hospital unless events were life threatening or required urgent care. More people dying at home may reflect this process,^26^ although the magnitude of increase is unlikely fully to explain our results especially the widening spatial inequalities. Similarly, COVID-19 and its legacy may have resulted in a permanent shock to the health system that has reduced the number of admitted care the NHS can provide. Alternatively, the lack of noticeable increases in avoidable hospitalisations may also suggest that the NHS and wider health system was resilient in coping with the pandemic, continuing to find ways to provide care in the community that prevented individuals being hospitalised for avoidable reasons. However, this should not be relied on given other evidence that the NHS is under “record pressures”.^27^

While trends in most of our outcome measures were broadly similar, vaccine-preventable hospital admissions were markedly different. Low levels of vaccine-preventable ambulatory care hospital admissions in 2020-2021 reflects the low level of influenza..^28^ Influenza normally accounts for a large proportion of these hospital admissions but, as with all respiratory infections, of its transmission is reduced by the non-pharmaceutical interventions adopted for SARS-CoV-2.^29,30^ Reductions in other viral and respiratory conditions such as asthma and COPD exacerbations also follow these trends.^24,31^ However, now that these measures have been relaxed, there is a need to prepare for co-occurring waves of SARS-CoV-2 and influenza.^32^

The impacts of the COVID-19 pandemic have in many respects been highly unequal.^8^ Individuals from socioeconomically deprived neighbourhoods, Black and Asian ethnic groups, and who reside in the north of England were more likely to experience severe outcomes related to COVID-19 including hospitalisation and mortality.^9,13,33^ These groups are often over-represented in unplanned hospital admissions and their greater needs for healthcare may have resulted in them being more affected by experiences of disruption.^16^ While our results provide evidence on the unequal distribution of avoidable hospitalisations across social and spatial characteristics, the changing trends in inequalities are not always so obvious, consistent and often nuanced in their interpretation.

Contrary to our hypothesis, we find evidence that absolute levels of inequalities in avoidable hospitalisations across deprivation, ethnicity and region largely narrowed in 2020 and 2021, particularly for deprivation. Counterintuitively, the narrowing of inequalities occurred during national lockdowns, suggesting that the periods of greatest disruption in the access to healthcare have not exacerbated inequalities. We cannot discount entirely the possibility that this is a consequence of fewer hospital admissions overall rather than the result of the many, often undocumented, responses taken by the NHS during the pandemic. It may also be that individuals who would have normally been captured in avoidable hospitalisations had experienced COVID-19, been hospitalised and died as a result instead (especially given the disproportionate distribution of unplanned hospitalisation and COVID-19 outcomes by our measures of inequality). However, 2021 and 2022 present differing trends by marker of inequalities with inconsistent or no trends for deprivation and ethnicity, but large increases in absolute and relative regional inequalities. This suggests that as we exit the pandemic, inequalities are becoming more entrenched geographically (particularly due to higher rates in northern regions). We are not aware of any other analysis which has investigated or found this, and our results demonstrate the need for identifying the reasons behind why place is becoming more important for describing inequalities in avoidable hospitalisations.

Although absolute inequalities did narrow, we still find evidence of differences between regions, ethnicity and levels of deprivation reminding us that inequalities remain important. More deprived areas and northern regions had consistently higher avoidable hospitalisations throughout the whole study period. There was a lack of literature on ethnic inequalities in avoidable hospitalisations pre-pandemic,^16,34^ and our work contributes to showing how intricate they can be. Our findings reiterate the need for policy makers to focus on tackling the root structural causes of health. Health is socially determined.^8^ While the unequal differences we describe are unjust, they are not inevitable and can be fixed. In particular, tackling social and spatial inequalities should be a core part of the UK Government’s ‘Levelling up’ agenda. While we do not find clear and consistent inequalities for ethnicity, this should not detract from the structural injustices that some ethnic group face which has implications for their health.^9,33^

### Limitations

Our analyses are descriptive and are unable to identify the reasons behind what the trends we observe. The findings may be subject to ecological fallacy as we had no measure of individual experience of healthcare disruption. However, the magnitude of changes are clear enough to suggest that they are real at least at the population level. While our electronic health records provide large representative data on patients,^18^ they lack the detail about the contexts of individual’s lives that can be found in other data types (e.g., longitudinal surveys). Future research should seek to link electronic health records to richer individual-level cohort data to be able to tease out the pathways and mechanisms that may explain our findings.

By using a proxy measure of health system performance, we are also unable to ascertain the extent that our findings do reflect actual disruption to healthcare. Its impacts will be complex, occurring over the short- and long-term. It may be that our study is too early to detect the full impact of this disruption and it will be necessary to continually monitor trends in our outcomes. Additionally, there is some debate in the literature over how valid avoidable hospitalisations are as a proxy for health system performance.^16^ Although our measures are used by NHS England for measuring health systm performance,^16^ other COVID-19 related outcomes have witnessed widening social inequalities which are different to our findings.^2,8,9^ Regional and hospital differences in reporting patterns may also exist. Validating our chosen outcomes, and comparing how their trends compare to other outcomes linked with capturing features of healthcare disruption, would be important for understanding the importance of our findings.

We use coarse groupings for each of our measures of inequality. For example, we group individuals by broad ethnic groups that hide the diversity within each group (e.g., Asian constitutes communities from very different backgrounds and heritages). This was partly to minimise statistical disclosure issues, due to the lower number of events per month in some groups. Future research should utilise finer groupings that can better describe the extent of inequalities, ideally recognising the important of intersectionality, and help identify particular populations at greater risk of avoidable hospitalisations. Similarly, moving beyond describing geographical inequalities by region to identify how trends vary across smaller places can help to present more precise patterns and help to identify potential drivers.

### Conclusions

The COVID-19 pandemic has affected avoidable hospitalisations in unexpected and potentially surprising ways. Worries that the pandemic would see rising hospital admissions due to disruptions in accessing care do not appear to have materialised, at least so far. While social and spatial inequalities narrowed during the pandemic, recent widening regional inequalities present cause for concern and present the case for renewed focus among narratives of ‘building back better’ and ‘levelling up’.

## Data Availability

All data were linked, stored and analysed securely within the OpenSAFELY platform: https://opensafely.org/. Data include pseudonymised data such as coded diagnoses, medications and physiological parameters. No free text data are included. All code is shared openly for review and re-use under MIT open license on https://github.com/opensafely/avoidable_hospitalisations_trends. Detailed pseudonymised patient data is potentially re-identifiable and therefore not shared. Primary care records managed by the GP software provider, TPP, were linked to Hospital Episode Statistics through OpenSAFELY.

https://github.com/opensafely/avoidable_hospitalisations_trends

## Acknowledgements

We are very grateful for all the support received from the TPP Technical Operations team throughout this work, and for generous assistance from the information governance and database teams at NHS England and the NHS England Transformation Directorate. Information about data sharing, data access and information governance can be found at the end of this document. This work was funded as part of the National Core Studies Longitudinal Health & Wellbeing programme (MC_PC_20030, MC_PC_20059). MAG acknowledges funding for this research from UK Medical Research Council (MR/W021242/1). SVK acknowledges funding from a NRS Senior Clinical Fellowship (SCAF/15/02), the Medical Research Council (MC_UU_00022/2) and the Scottish Government Chief Scientist Office (SPHSU17). JM is partly funded by the National Institute for Health and Care Research Applied Research Collaboration West (NIHR ARC West). The full statement, listing the names of all relevant NCS Consortium staff, can be found here: https://www.ucl.ac.uk/covid-19-longitudinal-health-wellbeing/convalescence-study-collaborative.

## Information governance

NHS England is the data controller for OpenSAFELY-TPP and OpenSAFELY-EMIS; EMIS and TPP are the data processors; all study authors using OpenSAFELY have the approval of NHS England. This implementation of OpenSAFELY is hosted within the EMIS and TPP environments which are accredited to the ISO 27001 information security standard and are NHS IG Toolkit compliant.^1^

Patient data has been pseudonymised for analysis and linkage using industry standard cryptographic hashing techniques; all pseudonymised datasets transmitted for linkage onto OpenSAFELY are encrypted; access to the platform is via a virtual private network (VPN) connection, restricted to a small group of researchers; the researchers hold contracts with NHS England and only access the platform to initiate database queries and statistical models; all database activity is logged; only aggregate statistical outputs leave the platform environment following best practice for anonymisation of results such as statistical disclosure control for low cell counts.^2^

The OpenSAFELY research platform adheres to the obligations of the UK General Data Protection Regulation (GDPR) and the Data Protection Act 2018. In March 2020, the Secretary of State for Health and Social Care used powers under the UK Health Service (Control of Patient Information) Regulations 2002 (COPI) to require organisations to process confidential patient information for the purposes of protecting public health, providing healthcare services to the public and monitoring and managing the COVID-19 outbreak and incidents of exposure; this sets aside the requirement for patient consent.^3^ This was extended in July 2022 for the NHS England OpenSAFELY COVID-19 research platform.^4^ In some cases of data sharing, the common law duty of confidence is met using, for example, patient consent or support from the Health Research Authority Confidentiality Advisory Group.^5^

Taken together, these provide the legal bases to link patient datasets on the OpenSAFELY platform. GP practices, from which the primary care data are obtained, are required to share relevant health information to support the public health response to the pandemic, and have been informed of the OpenSAFELY analytics platform.

This study was classified as service evaluation. This study was supported by Professor Martin McKee (Honorary Consultant at UCLH NHS Foundation Trust) as senior sponsor, and approved by the University of Liverpool’s Research Ethics Board [reference 10634].

## Data access

Access to the underlying identifiable and potentially re-identifiable pseudonymised electronic health record data is tightly governed by various legislative and regulatory frameworks, and restricted by best practice. The data in OpenSAFELY is drawn from General Practice data across England where EMIS and TPP are the data processors.

EMIS and TPP developers initiate an automated process to create pseudonymised records in the core OpenSAFELY database, which are copies of key structured data tables in the identifiable records. These pseudonymised records are linked onto key external data resources that have also been pseudonymised via SHA-512 one-way hashing of NHS numbers using a shared salt. Bennett Institute for Applied Data Science developers and PIs holding contracts with NHS England have access to the OpenSAFELY pseudonymised data tables as needed to develop the OpenSAFELY tools.

These tools in turn enable researchers with OpenSAFELY data access agreements to write and execute code for data management and data analysis without direct access to the underlying raw pseudonymised patient data, and to review the outputs of this code. All code for the full data management pipeline—from raw data to completed results for this analysis—and for the OpenSAFELY platform as a whole is available for review at github.com/OpenSAFELY.

The data management and analysis code for this paper was led by Dr Mark Green and contributed to by all named authors.

## Notes

### Competing Interest Statement

The authors have declared no competing interest.

### Author Declarations

This study was classified as service evaluation. This study was supported by Professor Martin McKee (Honorary Consultant at UCLH NHS Foundation Trust) as senior sponsor, and approved by the University of Liverpool Research Ethics Board [reference 10634].

