## appendix for "Trends in inequalities in avoidable hospitalisations across the COVID-19 pandemic: A cohort study of 23.5 million people in England"

**Table A: Missing data for covariates**

| Measure | Percent missing (%) |
| --- | --- |
| Index of Multiple Deprivation | 1.68 |
| Ethnicity | 10.21 |
| Government Office Region | 0.05 |

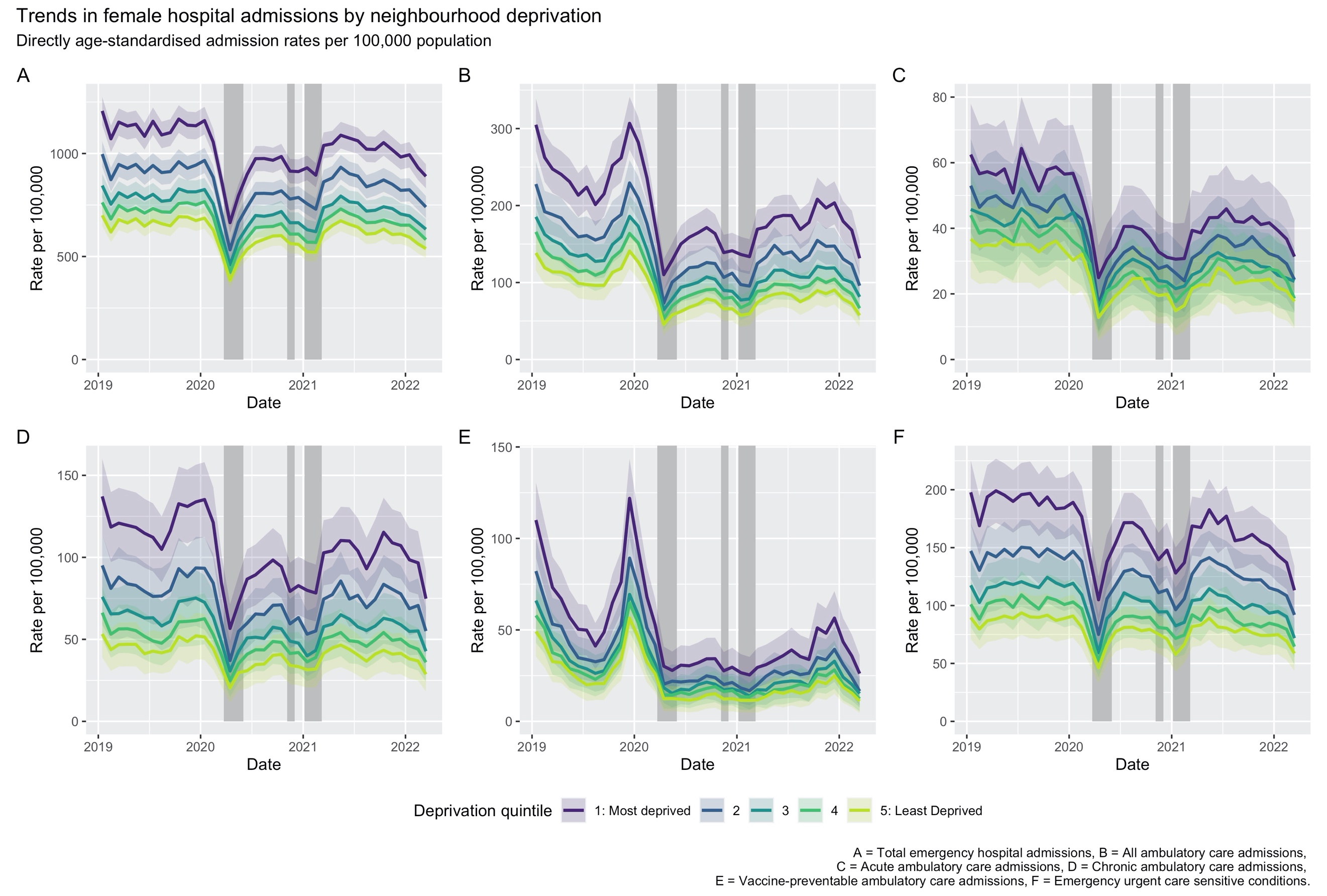

**Figure A: Directly age-standardised admission rates for females by socioeconomic deprivation quintile.**

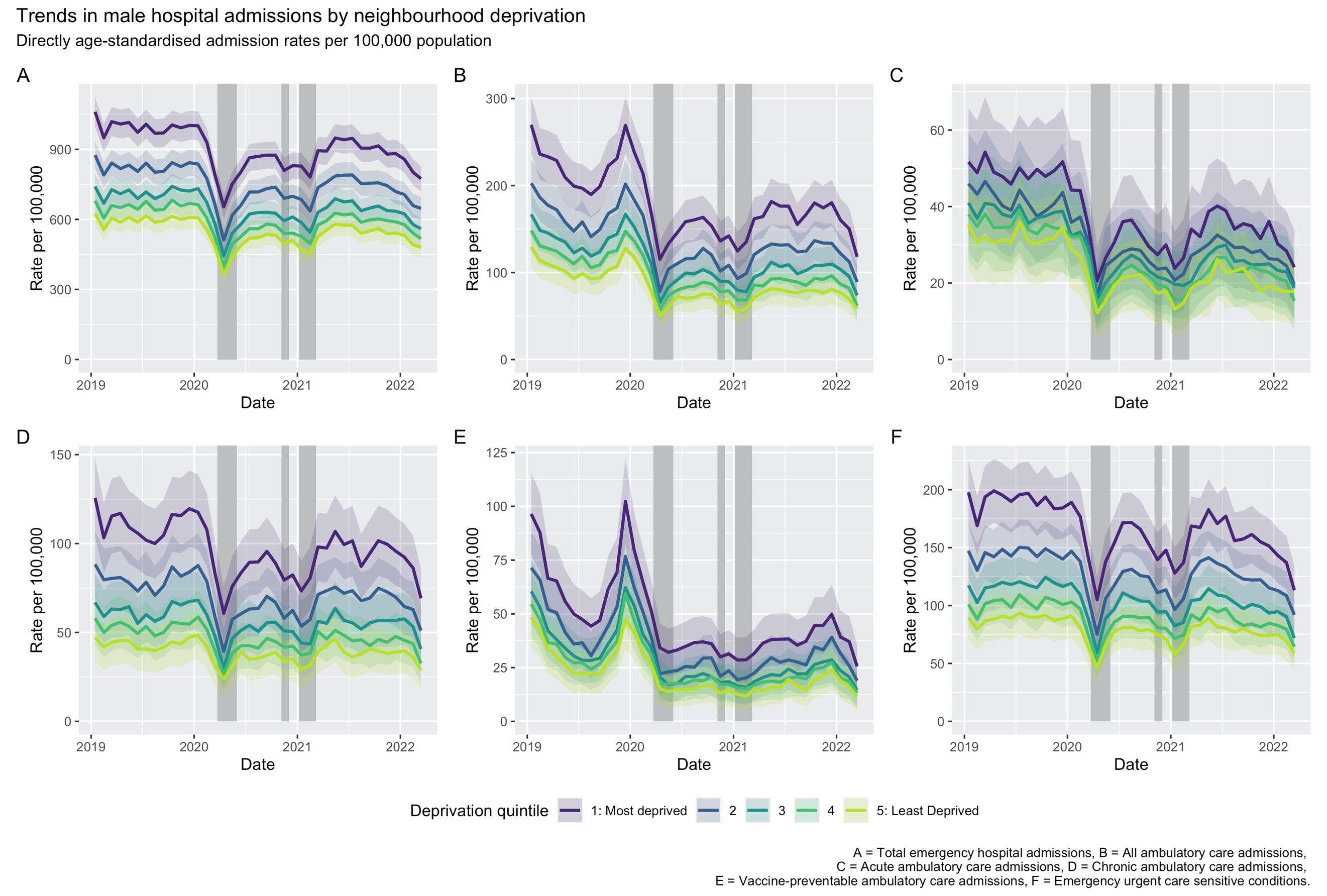

**Figure B: Directly age-standardised admission rates for males by socioeconomic deprivation quintile.**

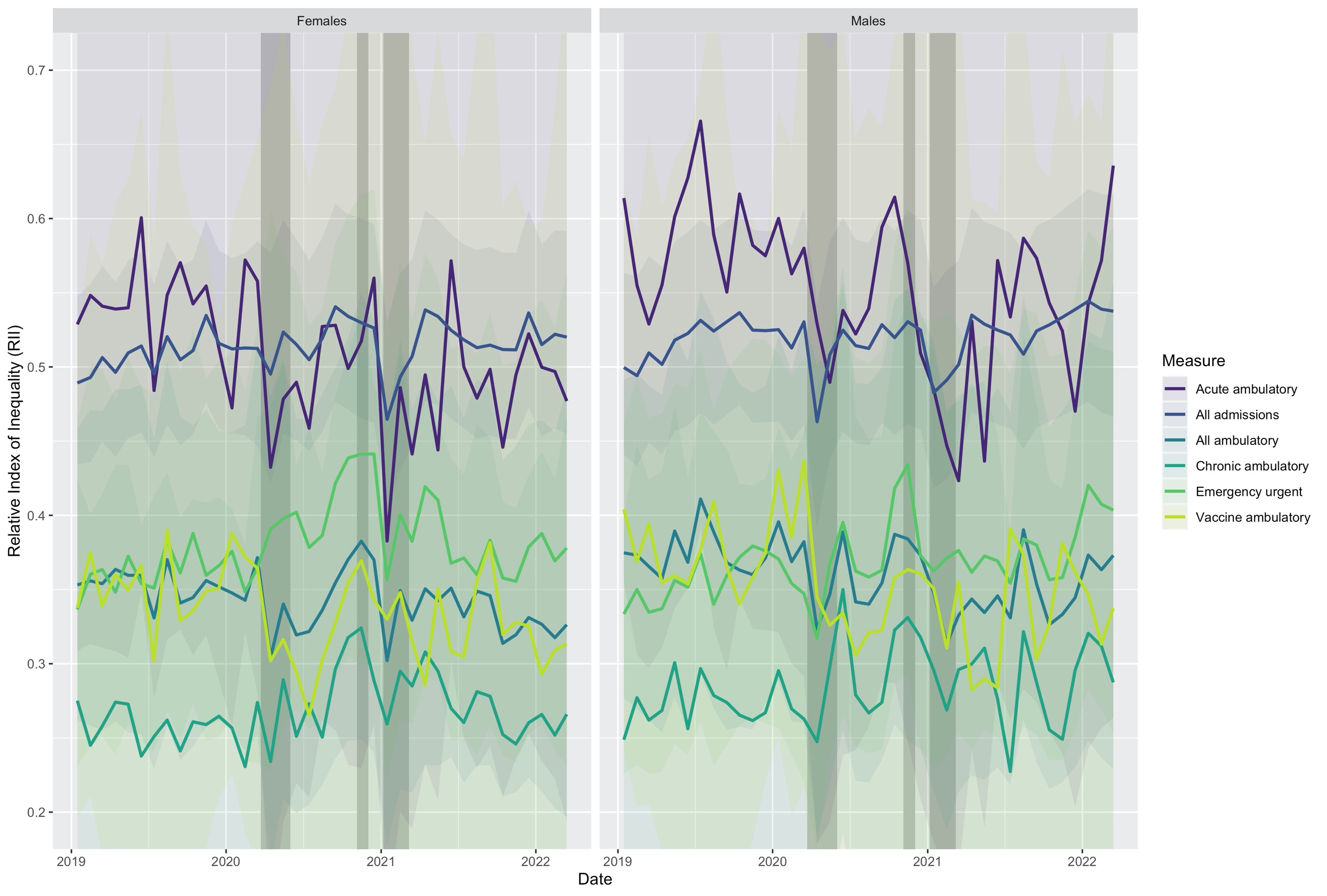

**Figure C: Estimated Relative Index of Inequality (RII) for socioeconomic deprivation by sex.**

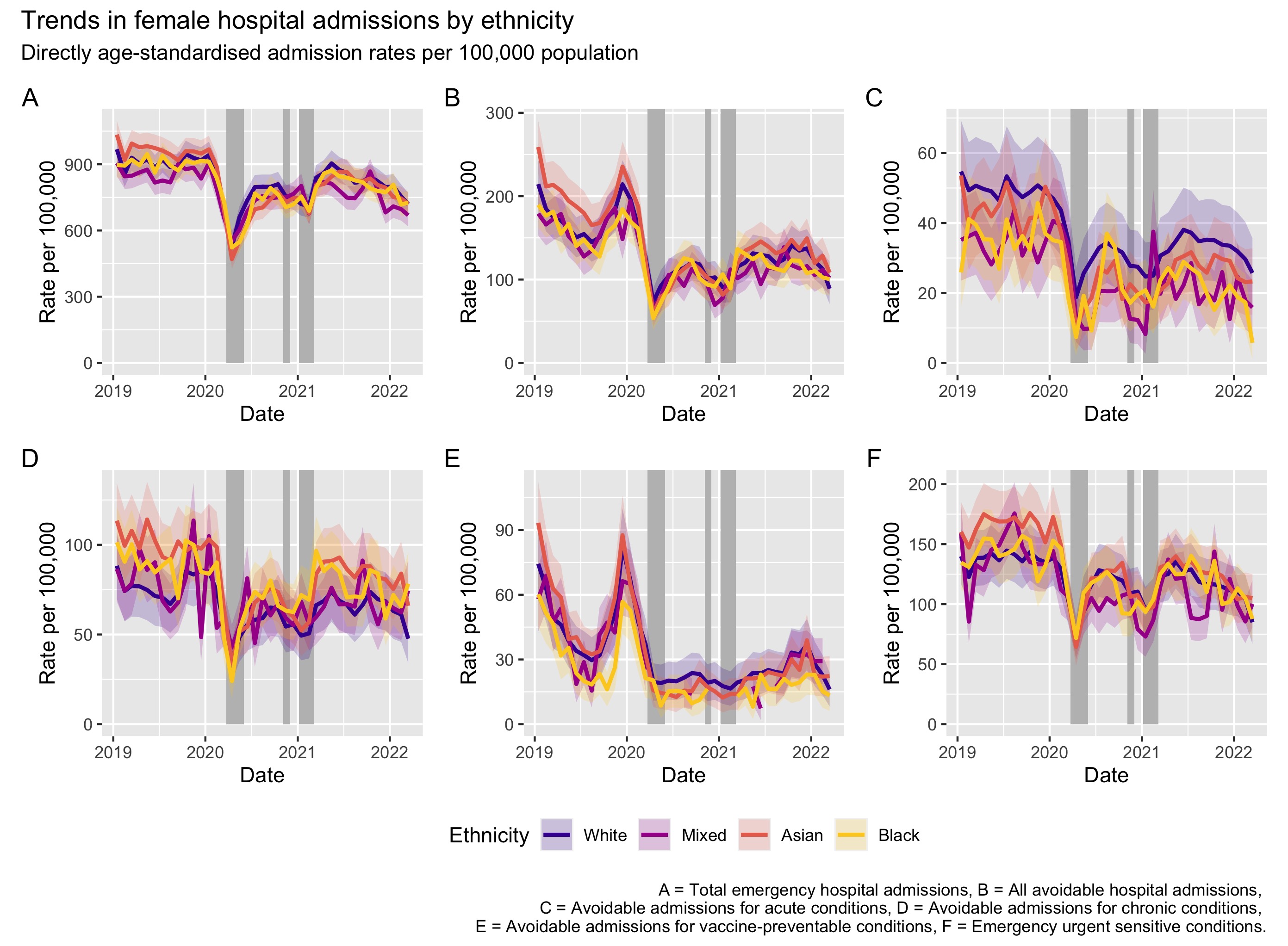

**Figure D:** **Directly age-standardised admission rates for females by ethnicity.**

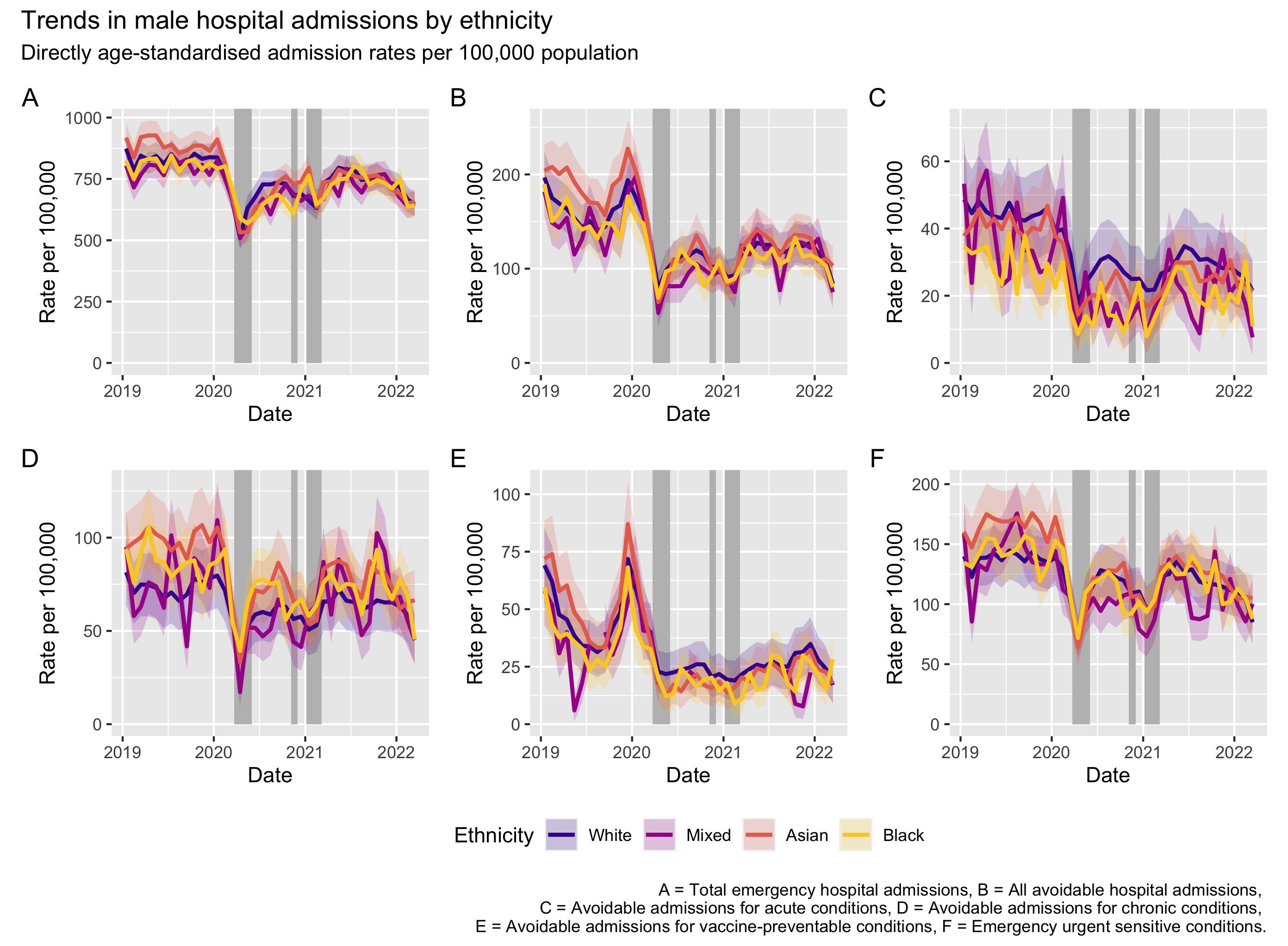

**Figure E:** **Directly age-standardised admission rates for males by ethnicity.**

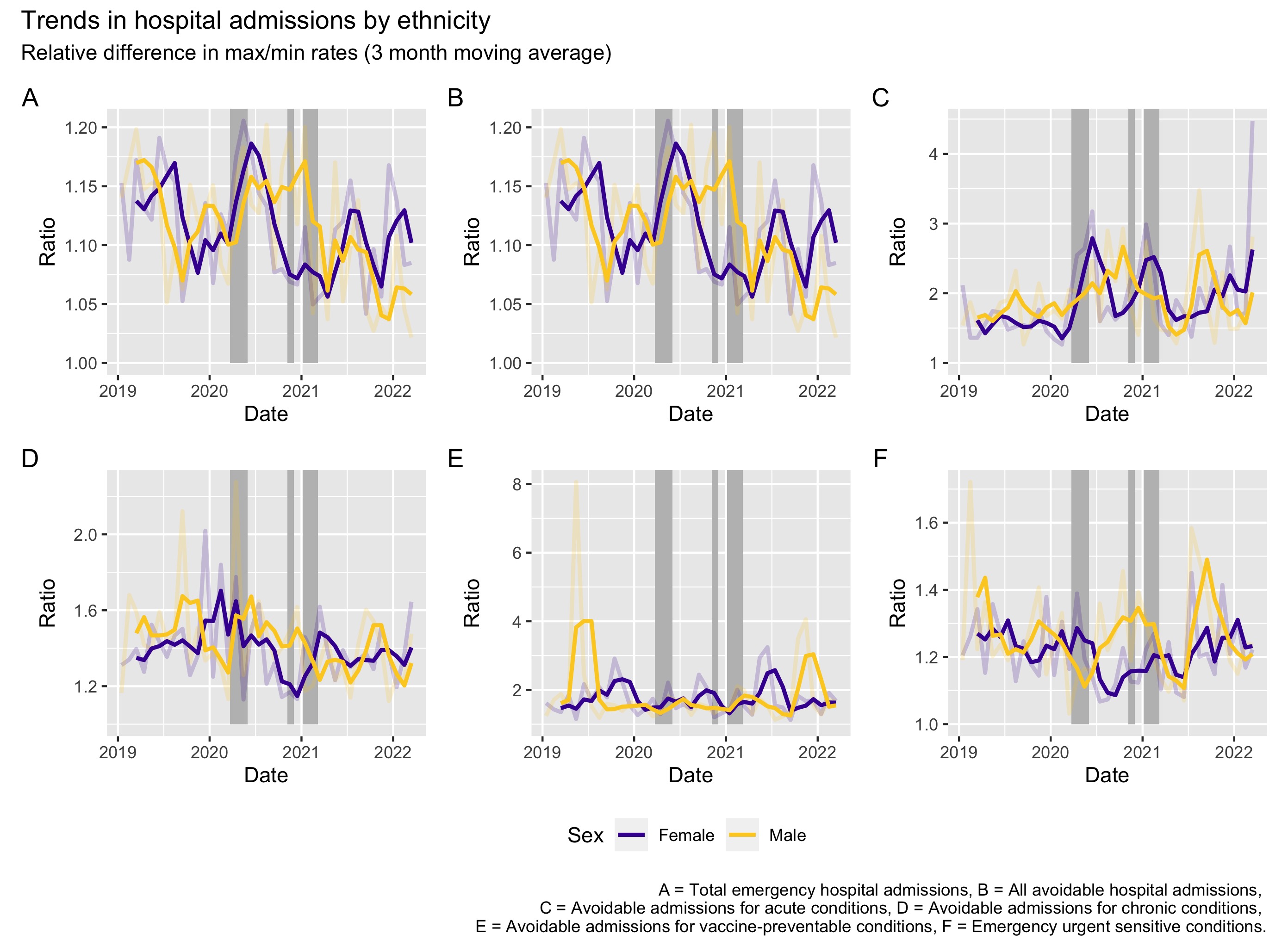

**Figure F: The ratio of directly standardised rates for admission (per 100,000) between the maximum and minimum values across ethnic groups and by cause.**

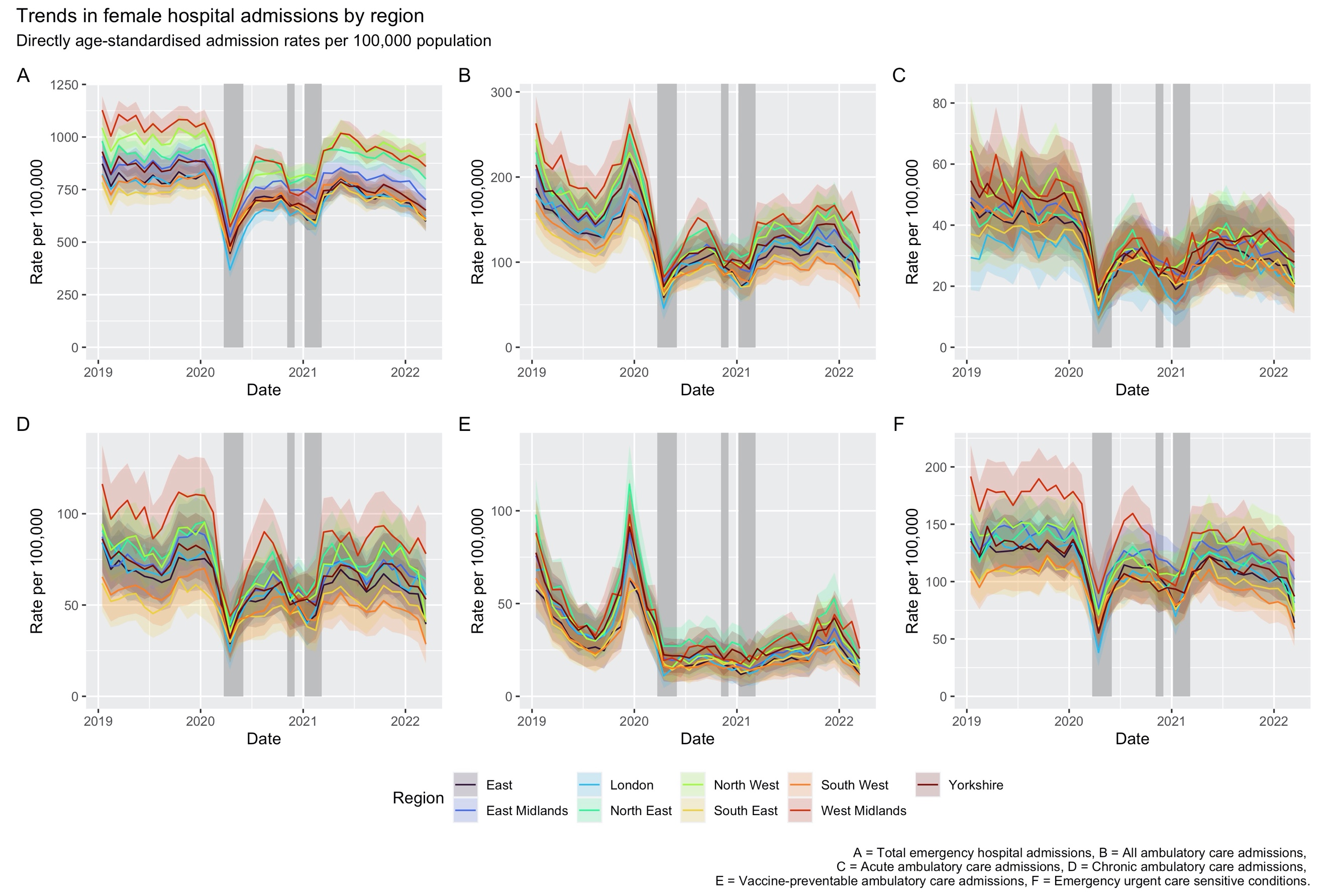

**Figure G:** **Directly age-standardised admission rates for females by region.**

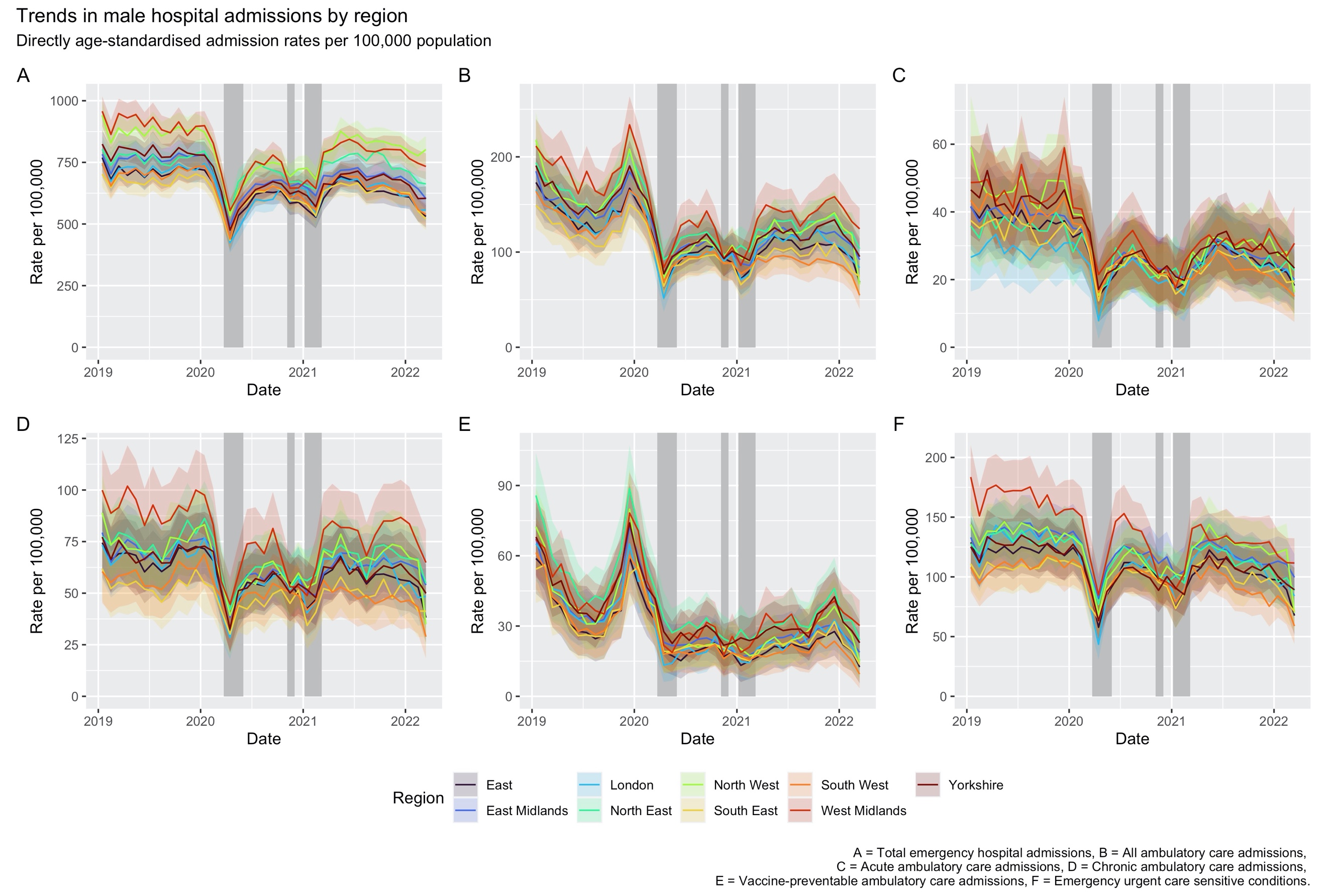

**Figure H:** **Directly age-standardised admission rates for males by region.**
